# QTNet: Predicting Drug-Induced QT prolongation with Deep Neural Networks

**DOI:** 10.1101/2021.03.24.21254235

**Authors:** Neil Jethani, Hao Zhang, Larry Chinitz, Yindalon Aphinyanaphongs, Rajesh Ranganath, Lior Jankelson

**Affiliations:** Department of Population Health, New York University Langone Health, New York, NY; Courant Institute of Mathematical Sciences, New York University, New York, NY; Leon H. Charney Division of Cardiology, Department of Medicine, New York University Langone Health, New York, NY

## Abstract

**Background:** Drug-induced QTc prolongation (diQTP) is frequent and associated with a risk of sudden cardiac death. Identifying patients at risk of diQTP can enhance monitoring and treatment plans.

**Objective:** To develop a machine learning architecture for prediction of extreme diQTP (QTc >500ms OR ΔQTc >60ms) at the onset of treatment with a QTc prolonging drug.

**Methods:** We included 4,628 adult patients who received a baseline ECG within 6 months prior to treatment onset with a QTc prolonging drug and a follow-up ECG after the fifth dose. We collected known clinical QTc prolongation risk factors (CF). We developed a novel neural network architecture (QTNet) to predict diQTP from both the CF and baseline ECG data (ECGD), composed of both the ECG waveform and measurements (i.e. QTc),by fusing a state-of-the-art convolution neural network to process raw ECG waveforms with the CF using amulti-layer perceptron. We fit a logistic regression model using the CF, replicating RISQ-PATH as Baseline. We further compared the performance of QTNet (CF+ECGD) to neural network models trained using three variable subsets: a) baseline QTc (QTC-NN), b) CF-NN, and c) ECGD-NN.

**Results:** diQTP was present in 1030 patients (22.3%), of which baseline QTc was normal (QTc<450ms:Male/<470ms:Female) in 405 patients (39.3%). QTNet achieved the best performance (Figure 1)(AUROC, 0.802 [95% CI, 0.782-0.820]), outperforming predictions based on the Baseline (AUROC, 0.738 [95%CI, 0.716-0.757]), QTC-NN (AUROC, 0.735 [95% CI, 0.710-0.757]), CF-NN (AUROC, 0.778 [95% CI, 0.757-0.799]), and ECGD-NN (AUROC, 0.774 [95% CI, 0.750-0.794]).

**Conclusion:** We developed QTNet, the first deep learning model for predicting extreme diQTP, outperforming models trained on known clinical risk factors.

## Research Letter

Drug-induced QTc prolongation (diQTP) is associated with increased risk of Torsades de Pointes and sudden cardiac death. Recently, widespread use of hydroxychloroquine as a treatment for COVID-19 put many patients at risk of these adverse events. Many more drugs are known to induce QTc prolongation, as listed by Crediblemeds^1^. We developed QTNet, a novel machine learning architecture which uses raw ECG waveforms, to predict the risk of developing QTc prolongation upon treatment onset with a QTc prolonging drug.

**Figure.**
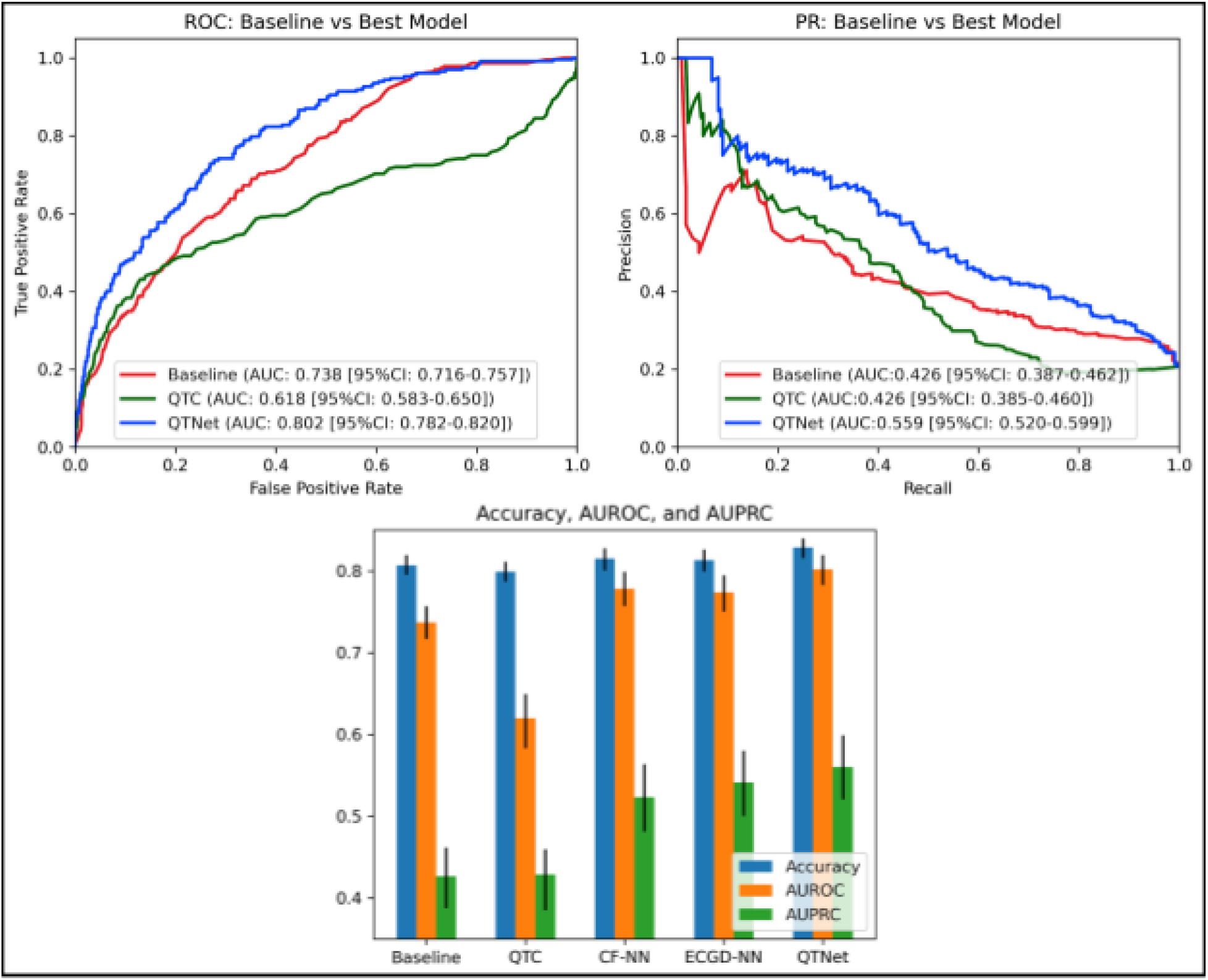
Performance of QTNet predicting diQTP. Compared models include the Baseline CF logistic regression model (Baseline), Reference QTc (QTC), NN trained on clinical features (CF-NN) and NN trained on ECG data (ECGD-NN). diQTP, drug induced QT prolongation; CF, clinical risk factors; NN, neural network.

We identified adults aged ≥ 18 years who were administered a QTc prolonging drug while admitted to New York University Langone Health between January 1, 2013 and January 13, 2021. A subset of the QT-prolonging medications with known risk for TdP, as defined by CredibleMeds^1^, were selected on the basis of administration frequency and expert opinion. This subset included Fluconazole, Hydroxychloroquine, Amiodarone, Donepezil, Escitalopram, Levofloxacin, Citalopram, Erythromycin, Ciprofloxacin, Haloperidol, and Dronedarone. Patients were eligible for inclusion if at least five doses of a QTc prolonging drug were administered, a “reference” 10s, 12 lead ECG was recorded within 6 months *prior* to the onset of treatment, and a “follow-up” ECG was recorded after the fifth dose and prior to drug cessation. Patients were split into training, validation, and test datasets in a 3:1:1 ratio.

Reference ECG data (ECGD) consisted of both the raw waveform and MUSE calculated measurements (i.e. PR interval). The corrected QT-interval (QTc) was calculated using the Bazett method. Extreme QTc prolongation, defined as QTc ≥ 500ms or ΔQTc≥ 60ms, present in any follow-up ECG was used as the study endpoint.

We collected known clinical QTc prolongation risk factors (CF) as identified in the RISQ-PATH^2^ model, which included patient demographics, laboratory values, vital signs, past medical history, and medication history as well the reference QTc measurement. Missingness indicators were included for each variable.

We developed a novel neural network architecture (QTNet) to predict diQTP from both the CF and ECGD, by fusing a state-of-the-art convolution neural network^3^ to process raw ECG waveforms with the CF using a fully-connected neural network. We tuned both the number of layers (1,2, or 3) and hidden dimension size (100, 1000, or 10000) of the multi-layer perceptron.

Applying the previously published RISQ-PATH model directly to our data performed poorly (AUROC, 0.537 [95% CI, 0.510-0.563]), therefore we refit a logistic regression model using the CF, replicating RISQ-PATH as Baseline. We also evaluated the use of the reference QTc (QTC) exclusively as a risk score.

We further compared the performance of QTNet (CF+ECGD) to neural networks (NN) trained using either the a) CF or b) ECGD. The validation dataset was used to select hyperparameters and checkpoints based on the area under the receiver-operator curve (AUROC). The results are reported using the hold-out test dataset.

A total of 4626 adult patients met the inclusion criteria. diQTP was present in 1030 patients (22.3%), of which reference QTc was normal (QTc <450ms for male, <470ms for female) in 405 patients (39.3%) prior to medication administration. The prevalence of QTc prolongation nor the patient risk factors and ECG measurements differed significantly across training, validation, and test set (p-values calculated using ANOVA for continuous and χ^2^ test for categorical variables and Bonferroni corrected for multiple hypotheses).

QTNet achieved the best performance (Figure 1) (AUROC, 0.802 [95% CI, 0.782-0.820]), outperforming predictions based on using the Baseline (AUROC, 0.738 [95% CI, 0.716-0.757]), QTC (AUROC, 0.620 [95% CI, 0.583-0.650]), CF-NN (AUROC, 0.778 [95% CI, 0.757-0.799]), and ECGD-NN (AUROC, 0.774 [95% CI, 0.750-0.794]). Of note, models trained with the CF and those trained with the ECGD alone appear to perform similarity across metrics.

Further, we performed the same analysis on additional endpoints, including moderate QTc prolongation (QTc ≥ 450ms for males, ≥ 470ms for females), JTc prolongation (JTc ≥ 350ms), and ΔJTc prolongation (ΔJTc ≥ 60) using both the Bazett and Frederica methods for calculating QTc and observed that QTNet was the best performing model.

Automated identification of patients at risk for diQTP is an important unmet need^4^. This is the first study to apply deep learning and suggest that reference ECG waveforms can be effectively used to predict diQTP risk, with a high AUROC of 0.802. Further, the performance of QTNet at 70% PPV (Sensitivity, 0.272 [95% CI, 0.093-0.401]) indicates that deployment of this model would allow many patients at high risk for diQTP to be identified. Contrary to rhythm detection, developing accurate deep learning models for complex clinical predictions present an ongoing challenge. QTNet may present a useful starting-point for fusing CF and ECGD to enhance complex clinical predictions.

## Data Availability

Data will not be available due to PHI inclusion

